# Cancer immunotherapy does not increase the risk of death by COVID-19 in melanoma patients

**DOI:** 10.1101/2020.05.19.20106971

**Authors:** Maria Gonzalez-Cao, Mónica Antoñanzas Basa, Teresa Puertolas, Eva Muñoz, José Luis Manzano, Cristina Carrera, Ivan Marquez-Rodas, Pilar Lopez Criado, Juan Francisco Rodriguez Moreno, Almudena García Castaño, Juan Martín-Liberal, Pedro Rodríguez-Jiménez, Susana Puig, Pablo Cerezuela, Marta Feito Rodríguez, Belén Rubio Viqueira, Guillermo Crespo, Pablo Luna Fra, Cristina Aguayo Zamora, Pablo Ayala de Miguel, Rosa Feltes Ochoa, Lara Valles, Ana Drozdowskyj, Ainara Soria, Cayetana Maldonado Seral, Luis Fernández-Morales, Rafael Rosell, Mariano Provencio, Alfonso Berrocal, for the Spanish Melanoma Group (GEM).

## Abstract

**Background:** Covid-19 pandemic by the new coronavirus SARS-Cov-2 has produced devastating effects on the health care system, affecting also cancer patient care. Data about COVID-19 infection in cancer patients are scarce, and they point out a higher risk of complications due to the viral infection in this population. Moreover, cancer treatments could increase viral complications, specially those treatments based on the use of immunotherapy with checkpoints antibodies. There are no clinical data about the safety of immune check point antibodies in cancer patients when they become infected by SARS-CoV-2. As checkpoint inhibitors, mainly anti PD-1 and anti CTLA-4 antibodies, are an effective treatment for most melanoma patients, avoiding their use during the pandemic could lead to a decrease in the chances of curing melanoma.

**Methods:** In Spain we have started a national registry of melanoma patients infected by SARS-Cov-2 since April 1^st^, 2020. A retrospective analysis of patients included in the Spanish registery has been performed weekly since the activation of the study. Interim analysis shows unexpected findings about cancer treatment safety in SARS-Cov-2 infected melanoma patients, so a rapid communication to the scientific community is mandatory

**Results:** Fifty patients have been included as of May 17^th^, 2020. Median age is 69 years (range 6 to 94 years), 27 (54%) patients are males and 36 (70%) patients have stage IV melanoma. Twenty-two (44%) patients were on active anticancer treatment with anti PD-1 antibodies, 16 (32%) patients were on treatment with BRAF plus MEK inhibitors and 12 (24%) patients were not on active cancer treatment. COVID-19 episode has been resolved in 43 cases, including 30 (70%) patients cured, four (9%) patients that have died due to melanoma progression, and nine (21%) patients that have died from COVID-19. Mortality rates from COVID-19 according to melanoma treatment type were 16%, 15% and 36% for patients on immunotherapy, targeted drugs, and for those that were not undergoing active cancer treatment, respectively.

**Conclusion:** These preliminary findings show that the risk of death in those patients under going treatment with anti PD-1 antibodies does not exceed the global risk of death in this population. These results could be relevant in order to select melanoma therapy during the COVID-19 pandemic

## Introduction

Data about COVID-19 evolution in cancer patients are scarce, but they point out a higher risk of severe events in this group^1-4^. Particularly, there are concerns in the scientific community about the safety of anti-cancer drugs and how they could interfere with aggressiveness of the viral infection, mainly immunotherapy with checkpoint inhibitors^5^. The largest study published until now about COVID-19 in cancer patients has included data from 105 patients, including only six patients treated with immunotherapy that had a COVID-19 mortality rate of 33%^6^. Antibodies against checkpoint inhibitors in T-cells, mainly PD-1 or CTLA-4, reverse the effects of the so-called exhausted phenotype of T-cells and have demonstrated high anti-tumoral activity in several malignancies, including melanoma^7^. Actually immunotherapy is one of the mainstays of melanoma treatment^8-12^. However, there are theoretical concerns about whether reinvigoration of immune response caused by these drugs could increase the risk of developing severe complications in case of SARS-Cov-2 viral infection^13^. This has led some physicians to take precautions about its use in melanoma, and recommendations have been made on the use of other therapeutic alternatives during the COVID-19 pandemic. As far as we know, there are no clinical data about the safety of these drugs in melanoma patients during the SARS-CoV-2 pandemic. Here we present the preliminary results of the Spanish Melanoma Register of COVID-19, analyzing the COVID-19 mortality rates according to cancer treatment type.

## Methods

The Spanish Melanoma Group (GEM) has started a national registry of melanoma patients infected by SARS-Cov-2. Patients previously diagnosed with melanoma, any stage, and with a clinical or RT-PCR confirmed infection of SARS-CoV-2, were included. By May 17th, 50 cases have been collected from 26 hospitals across the country. A weekly retrospective analysis has been performed since April 1^st^, 2020. Every analysis has included a description of patients characteristics, demographic and clinical features, cancer treatment (received in the last eight weeks), as well as mortality by COVID-19 or other causes. Categorical variables are summarized reporting the number (n) and percentage (%) of patients on each category and group. Continuous variables are summarized repoprting the sample size, median and range, on each group. Differences of clinical chracteristics between treatment subgroups where analyzed by using Chisquare test or Fisher exact test, as appropiated on each case, for categorical variables and the non-parametric Wilcoxon rank-sum test for continuous variables. The interim analysis of the first fifty patients registered is communicated in this manuscript. This study was approved on March 23^rd^, 2020 by the Ethics Committee of the Hospital Puerta del Hierro, Madrid (Exp. 6/2020). The requirement for patient informed consent was waived by the Ethics Commitee due to the emergency research circumstances.

## Results

Fifty patients have been included through May 17^th^, 2020. Median age is 69 years (range 6 to 94 years), 27 (54%) patients are males and 36 (70%) patients have stage IV melanoma (Table 1). Thirty-eight patients were undergoing active anti-cancer treatment including 22 (44%) patients treated with anti PD-1 antibodies and 16 (32%) were in treatment with BRAF plus MEK inhibitors. Twelve (24%) patients were not undergoing active cancer treatment (10 patients because they had a previous melanoma stage I-III surgically treated, one patient because he had a stage IV melanoma in complete response to previous immunotherapy that he had received three years before and the last patient because she was an advanced case on palliative care). From the total of patients included, only two cases (4%) were detected as being asymptomatic, since the screening of asymptomatic patients is not usually performed in Spain. Thirteen (26%) cases had mild COVID-19 symptoms, that did not require hospitalization, 23 (46%) patients developed pneumonia with severe symptoms, and twelve (24%) had life threatening complications. Eleven (22%) cases were outpatient managed, from the remaining cases, only four (8%) were ICU admitted. Until now, COVID-19 episode has been resolved in 43 cases, including 30 (70%) patients cured, four (9%) patients that have died due to melanoma progression, and nine (21%) patients that have died from COVID-19. As it was expected, most patients that died of COVID-19 were males (79% versus 42% in patients with COVID-19 resolved, p=0.0648) and were older than cured patients (median age was 78 years old -range 70 to 91 years-versus 65 years old -range 6 to 94 years-, p=0.0079). Mortality rate by COVID-19 was 23% and 20% in localized stages and stage IV melanoma, respectively.

According to therapy, mortality rates from COVID-19 were 16%, 15% and 36% for immunotherapy, targeted drugs, and for patients that were not undergoing active cancer treatment (p=0.7348) (Figure 1). Patient characteristics according to treatment type demonstrated that most patients without active anti cancer treatment were males (75%) (p=0.0941), had non metastatic melanoma (83%) (p <0.0001) and were outpatient treated (p=0.0206) (Table 1).

**Figure 1.**
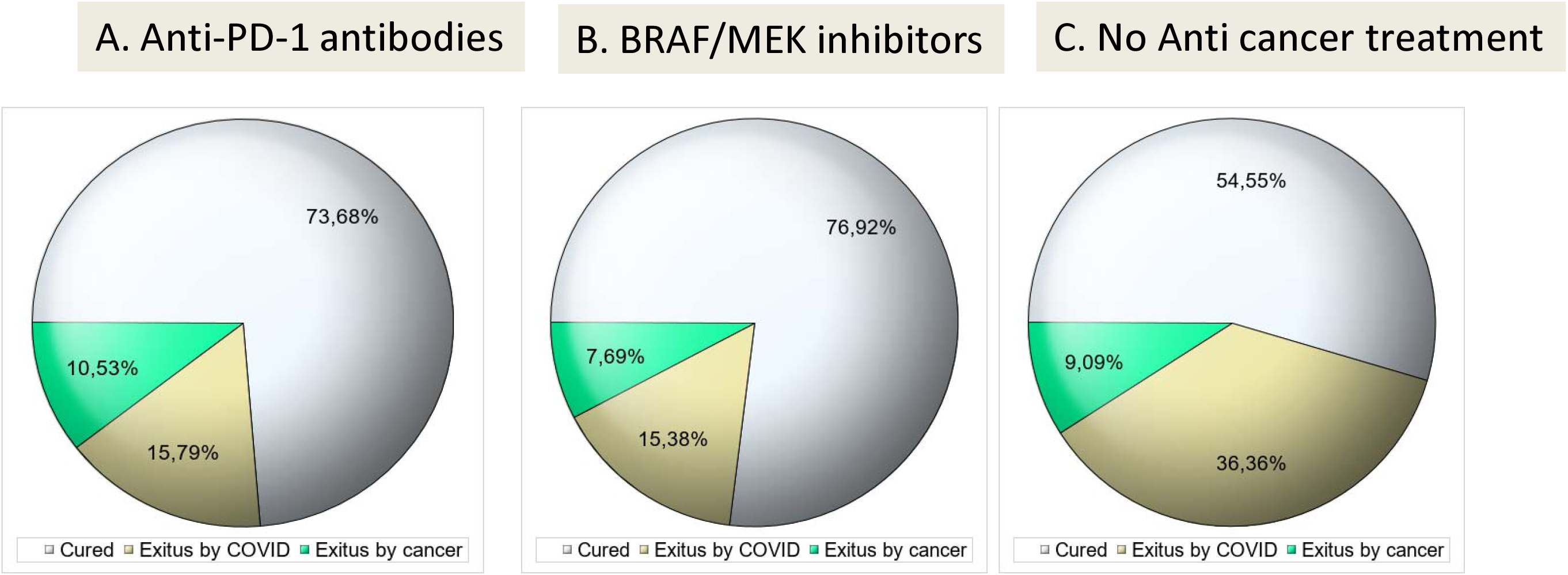
-Mortality and cure rates of SARS-Cov-2 infection in melanoma patients according to cancer treatment type

**Table 1.**
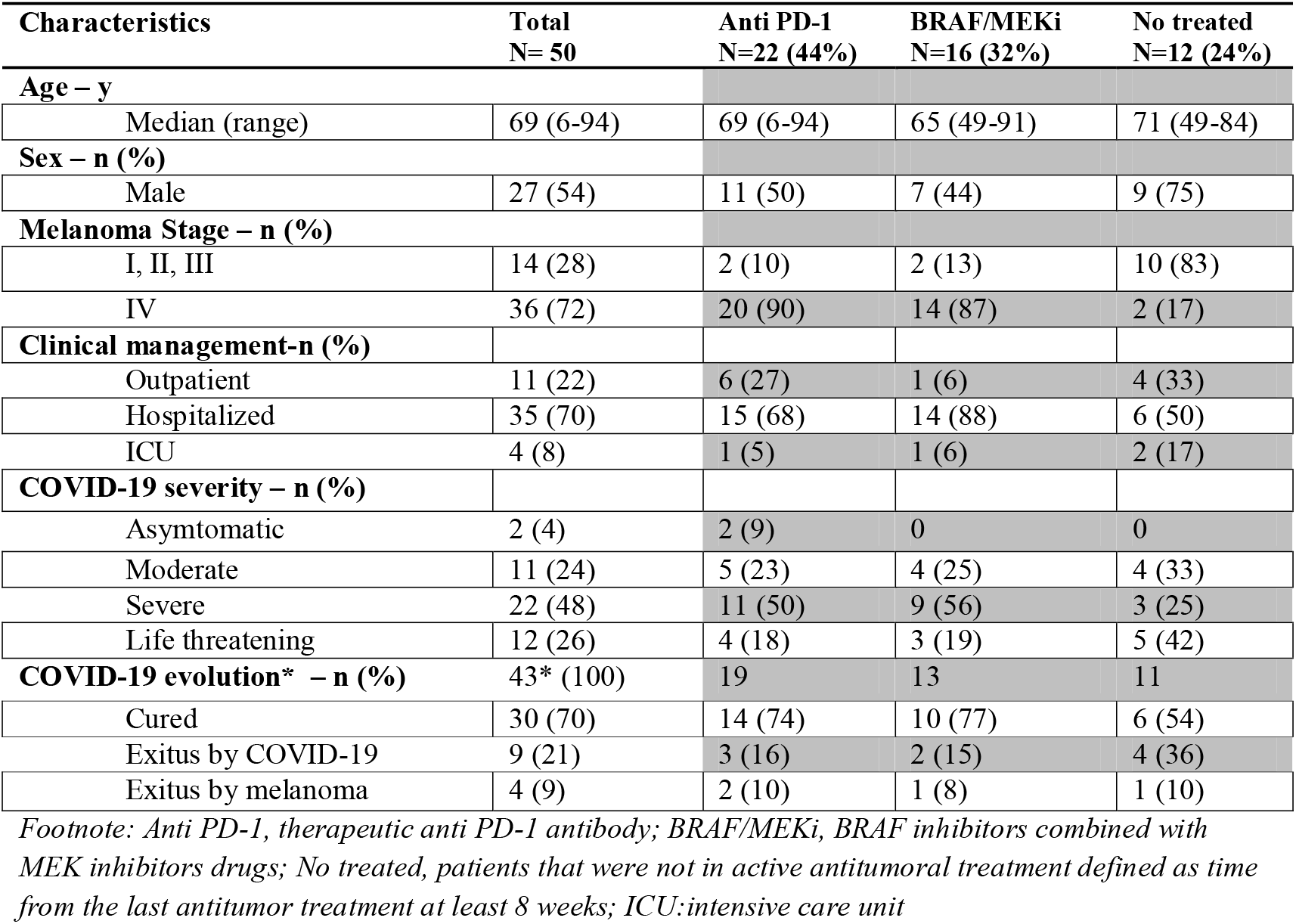
Baseline characteristics of COVID-19 infected melanoma patients

| Characteristics | Total<br>N= 50 | Anti PD-1<br>N=22 (44%) | BRAF/MEKi<br>N=16 (32%) | No treated<br>N=12 (24%) |
| --- | --- | --- | --- | --- |
| **Age – y** |  |  |  |  |
| Median (range) | 69 (6-94) | 69 (6-94) | 65 (49-91) | 71 (49-84) |
| **Sex – n (%)** |  |  |  |  |
| Male | 27 (54) | 11 (50) | 7 (44) | 9 (75) |
| **Melanoma Stage – n (%)** |  |  |  |  |
| I, II, III | 14 (28) | 2 (10) | 2 (13) | 10 (83) |
| IV | 36 (72) | 20 (90) | 14 (87) | 2 (17) |
| **Clinical management-n (%)** |  |  |  |  |
| Outpatient | 11 (22) | 6 (27) | 1 (6) | 4 (33) |
| Hospitalized | 35 (70) | 15 (68) | 14 (88) | 6 (50) |
| ICU | 4 (8) | 1 (5) | 1 (6) | 2 (17) |
| **COVID-19 severity – n (%)** |  |  |  |  |
| Asymptomatic | 2 (4) | 2 (9) | 0 | 0 |
| Moderate | 11 (24) | 5 (23) | 4 (25) | 4 (33) |
| Severe | 22 (48) | 11 (50) | 9 (56) | 3 (25) |
| Life threatening | 12 (26) | 4 (18) | 3 (19) | 5 (42) |
| **COVID-19 evolution\* – n (%)** | 43* (100) | 19 | 13 | 11 |
| Cured | 30 (70) | 14 (74) | 10 (77) | 6 (54) |
| Exitus by COVID-19 | 9 (21) | 3 (16) | 2 (15) | 4 (36) |
| Exitus by melanoma | 4 (9) | 2 (10) | 1 (8) | 1 (10) |

Footnote: Anti PD-1, therapeutic anti PD-1 antibody; BRAF/MEKi, BRAF inhibitors combined with MEK inhibitors drugs; No treated, patients that were not in active antitumoral treatment defined as time from the last antitumor treatment at least 8 weeks; ICU: intensive care unit

## Discussion

Mortality rate in melanoma patients treated with immunotherapy does not exceed the global mortality rate in this population, according to our data. The clinical implications of this finding could be relevant, as treatment of melanoma patients with checkpoint inhibitors during the COVID-19 pandemic has been avoided by many specialists. Immunotherapy with check point inhibitors, mainly anti PD-1 antibodies, can produce long lasting remissions in advanced melanoma patients^7,12,13^, as well as cures, when they are used as adjuvant therapy after surgery of localized melanoma.^14^ Therefore, avoiding the use of checkpoint inhibitors could have severe consequences in melanoma control. Theoretical concerns about the possibility of anti PD-1 antibodies having detrimental effects on COVID-19 evolution is mainly based on the fact that the pathophysiology of COVID-19 complications resembles that of immune related toxicity by anti-cancer checkpoint inhibitors^13,15-17^. SARS-Cov-2 infection, as it happens with toxic effects of anti PD-1 or anti CTLA-4 antibodies, may lead to an excessive host immune response that could produce an immune related pneumonitis with a high infiltration of the lung tissues by macrophages and activated T-cells, with a cytokine storm syndrome that ultimately leads to fatal multi-organ damage^15,16^. On the other hand, activated virus specific T-cells are also necessary for controlling the viral infection, while an increase of exhausted T-cells with high PD-1 expression is linked to severe COVID-19 evolution^18^. The cure of COVID-19 patients needs a delicate balance between immune response activation and control of excessive activation of the host immune response. Some investigators of different countries have designed clinical trials for analyzing the antiviral activity of anti-PD-1 antibodies in COVID-19 patients (NCT04356508, NCT04268537, NCT04335305), but data are pending and close monitoring of potential toxic effects will be necessary.

Other studies on COVID-19 evolution in cancer patients have included patients with a heterogeneous cancer types, most had lung cancer or gastrointestinal tumors, but none included melanoma patients^2,3,5,17^. These studies have found a high rate of severe complications from SARS-CoV-2 infection, 54%^3^, with a mortality rate of 18%-28.6%^2,3,17^of the whole population. Such results are in accordance with the preliminary findings of our analysis that show a rate of severe complications in 48% of patients and a mortality rate of 21%.

The unexpected findings of our analysis deserve special mention, showing a high mortality rate in those patients without active anti-cancer treatment. In this subgroup of twelve patients, most cancer free with localized melanoma stages, the COVID-19 mortality rate was 36%, while in patients undergoing active therapy the mortality rate was 15%-16%, despite the fact that most patients undergoing active anti-cancer treatment had metastatic melanoma. Other studies including patients with localized stages of cancer have demonstrated opposite results, with an inferior risk of severe complications and death by COVID-19 in patients with non-metastatic tumors, compared to advanced stages^3,6^. This discrepancy could be due to the low number of patients included in our analysis, but other factors could also explain the higher mortality rate found in melanoma patients without anti-cancer treatment. Most patients that had no active anti-cancer treatment from our registry were males (75%) and were older than patients undergoing active anti-cancer treatment. Furthermore, some antiviral effects of anticancer treatments on SARS-CoV-2 infection cannot be ruled out, as well as the coexistence of other co-morbidities that could interfere with these results.

In conclusion, although further research is needed in order to clarify these findings, the results demonstrate that cancer immunotherapy with anti PD-1 antibodies does not increase the risk of complications by COVID-19 when melanoma patients become infected. These results support the fact that melanoma immunotherapy must not be avoided during the COVID-19 pandemic and patients must have access to cancer treatment during this new era.

## Data Availability

Database and analysis is available for the Spanish Melanoma Group

## Notes

### Competing Interest Statement

The authors have declared no competing interest.

### Funding Statement

No external funding has been received

